# Traumatic subarachnoid hemorrhage: a scoping review

**DOI:** 10.1101/2021.01.10.21249416

**Authors:** Dylan P Griswold, Laura Fernandez, Andres M Rubiano

## Abstract

Sixty-nine million people suffer from traumatic brain injury (TBI) each year, and TBI is the most common cause of subarachnoid hemorrhage (SAH). Traumatic SAH (tSAH) has been described as an adverse prognostic factor leading to progressive neurological deterioration and increased morbidity and mortality. However, a limited number of studies evaluate recent trends in the diagnostic and management of SAH in the context of trauma. The objective of this scoping review was to understand the extent and type of evidence concerning the diagnostic criteria and management of traumatic subarachnoid hemorrhage. This scoping review was conducted following the JBI methodology for scoping reviews. The review included adults who suffered SAH secondary to trauma. Data extracted from each study included study aim, country, methodology, population characteristics, outcome measures, a summary of findings, and future directives. Thirty studies met inclusion criteria. Studies were grouped into five categories by topic: tSAH associated with mild TBI (n=13), and severe TBI (n=3); clinical management and diagnosis (n=9); imaging (n=3); and 5) aneurysmal tSAH (n=1). Of the 30 studies, two came from a low-and middle-income country (LMIC); excluding China, nearly a high-income country. Patients with tSAH associated with mTBI have a very low risk of clinical deterioration and surgical intervention and should be managed conservatively when considering ICU admission. The Helsinki and Stockholm CT scoring systems, in addition to the AIS, Cr, age decision tree, may be valuable tools to use when predicting outcome and mortality.

## Introduction

It is estimated that, globally, 69 million people (95% CI 64–74 million) suffer from traumatic brain injury (TBI) each year.^1^ High-income countries (HICs) have nearly 18 million cases, while low-and middle-income countries (LMICs) have around 50 million cases - an almost three-fold increase.^1^ This is relevant because TBI is the most common cause of subarachnoid hemorrhage (SAH). Thus, traumatic subarachnoid hemorrhage (TSAH) is a common finding in moderate and severe TBI (sTBI), as it occurs in 33-60% of patients.^2,3^ Road traffic accidents, falls, and violence are the main contributing factors to sTBI, and the majority of victims are those of 15 to 44 years old in the prime of life and leading contributors to the country’s gross domestic product (GDP). Thus, a country’s economic security is affected by sTBI and should have a vested interest in reducing its prevalence.^4^

Although it is necessary to understand this condition’s pathophysiology more fully, some theories have been described in animal studies that could largely explain the clinical course of TSAH. These theories are principally concerned with the phenomenon of traumatic vasoconstriction, which contributes to secondary ischemic damage and has a variable incidence range of 19-68%. Marmarou et al. and Thomas et al. used a rat model to describe the significant increase of intracranial pressure (ICP) and mean arterial blood pressure changes, which occur as a compensatory mechanism to maintain normal cerebral perfusion pressure.^2,5,6^

TSAH has been described as an adverse prognostic factor leading to progressive neurological deterioration and increased morbidity and mortality. This is due to its related events of vasospasm, dyselectrolytemia, pituitary dysfunction, hypoxia, intracranial hypertension, and hydrocephalus.^3^

Current resources aim to understand the diagnosis and treatment of patients with SAH according to the severity degree of the trauma. The goal is to use this information to evaluate the cost-effectiveness of current management, reduce the length of stay, and redirect the use of already limited resources.^7^ Recently published studies have mentioned that patients with SAH secondary to mild TBI have a lower risk of clinical deterioration and surgical intervention,^8^ whereby the routine implementation of CT scans, mandatory neurosurgery consultations, and high-intensity observations are not necessary in most cases.^7,9^

TSAH is a public health problem of significant proportions due to the global burden of disease and its disproportionate effect on LMICs. While research has made it possible to improve the use of resource-stratified clinical interventions, it is not enough.^7^ Economies are dependent on fiscally active adults, and TSAH stunts the growth of GDP in LMICs. The implications, then, lie beyond the scope of medicine and must be taken up by economists and politicians.

Therefore, the objective of this scoping review is to develop a better understanding of TSAH. This scoping review will serve as an initial step in providing more evidence for healthcare professionals, economists, and policymakers in order that they might devote more resources towards this significant problem affecting both health and economic outcomes worldwide.

A preliminary search of MEDLINE, the Cochrane Database of Systematic Reviews, and JBI Evidence Synthesis was conducted, and no current or underway systematic reviews or scoping reviews on the topic were identified.

## Methods

The proposed scoping review was conducted following the JBI methodology for scoping reviews.^10^

### Inclusion criteria

#### Participants

Studies of adult and adolescent (>15 years old) patients were included. All studies of pediatric patients (<15 years old) were excluded.

#### Concept

The concept of interest for this scoping review were studies of subarachnoid hemorrhage secondary to traumatic brain injury. All studies focused on non-traumatic subarachnoid hemorrhage were excluded.

#### Context

The review was limited to studies conducted between 2005-2020.

#### Search strategy

The search strategy aimed to locate both published and unpublished studies. An initial limited search of MEDLINE and Scopus was undertaken to identify articles on the topic. The text words in the titles and abstracts of relevant articles, and the index terms used to describe the articles were used to develop a full search strategy for PubMed, MEDLINE, and Scopus. A full search strategy for MEDLINE is detailed in Appendix I. In the second phase of the search, the search strategy was adopted to search EMBASE, Web of Science, and EBSCO. The reference lists of selected studies were screened for additional studies during the third phase of the search.

Studies published in English, Spanish, and French between the years 2005-2020 were included.

#### Information sources

The databases searched included PubMed, Scopus, Embase, Web of Science, EBSCO, and MEDLINE.

#### Types of Sources

This scoping review considered experimental and quasi-experimental study designs, including randomized controlled trials, non-randomized controlled trials, before and after studies, and interrupted time-series studies. Analytical observational studies, including prospective and retrospective cohort studies, case-control studies, and analytical cross-sectional studies, were considered inclusion. This review also considered descriptive observational study designs, including case series, individual case reports, and descriptive cross-sectional studies for inclusion. Also, systematic reviews that met the inclusion criteria were considered. Text and opinion papers were also considered for inclusion in this scoping review.

#### Study/Source of Evidence selection

Following the search, all identified citations were collated and uploaded into EndNoteX9 (Clarivate Analytics, PA, USA). The citations were then imported into Covidence online software (Veritas Health Innovation, Melbourne, Australia) for screening. Two independent researchers (DG and LF) examined titles and abstracts for inclusion. The full texts of selected studies were retrieved and assessed. Full-text studies that did not meet the inclusion criteria were excluded, and the reasons for exclusion are listed in the Preferred Reporting Items for Systematic Reviews and Meta-Analyses (PRISMA) flow diagram. Any disagreements that arose between the researchers during either title and abstract screening or full-text screening were resolved through discussion. Included studies underwent a process of data extraction.

The results of the search are presented in the PRISMA flow diagram.

#### Data extraction

Data were extracted from the included studies by a reviewer and verified by a second reviewer using a data extraction tool developed in Covidence.

The data extracted included study aim, country, methodology, length of study, sample size, population characteristics, outcome measures, a summary of findings, and conclusion and future directives. Extracted data is available as a supplement.

#### Data synthesis

Studies were summarized in tables, graphs, and narratively.

## Results

### Study inclusion

The study selection process is illustrated in the PRISMA flow diagram in Figure 1.^11^The described search identified 2,978 records to screen, after which were left 81 studies for full-text review. A total of 52 articles were excluded (Supplement 1. Table 1). Reasons for exclusion were: wrong study design (n=27), wrong outcomes (n=14), wrong patient population (n=10), wrong language (n=1). This left 30 studies for inclusion in the final synthesis.

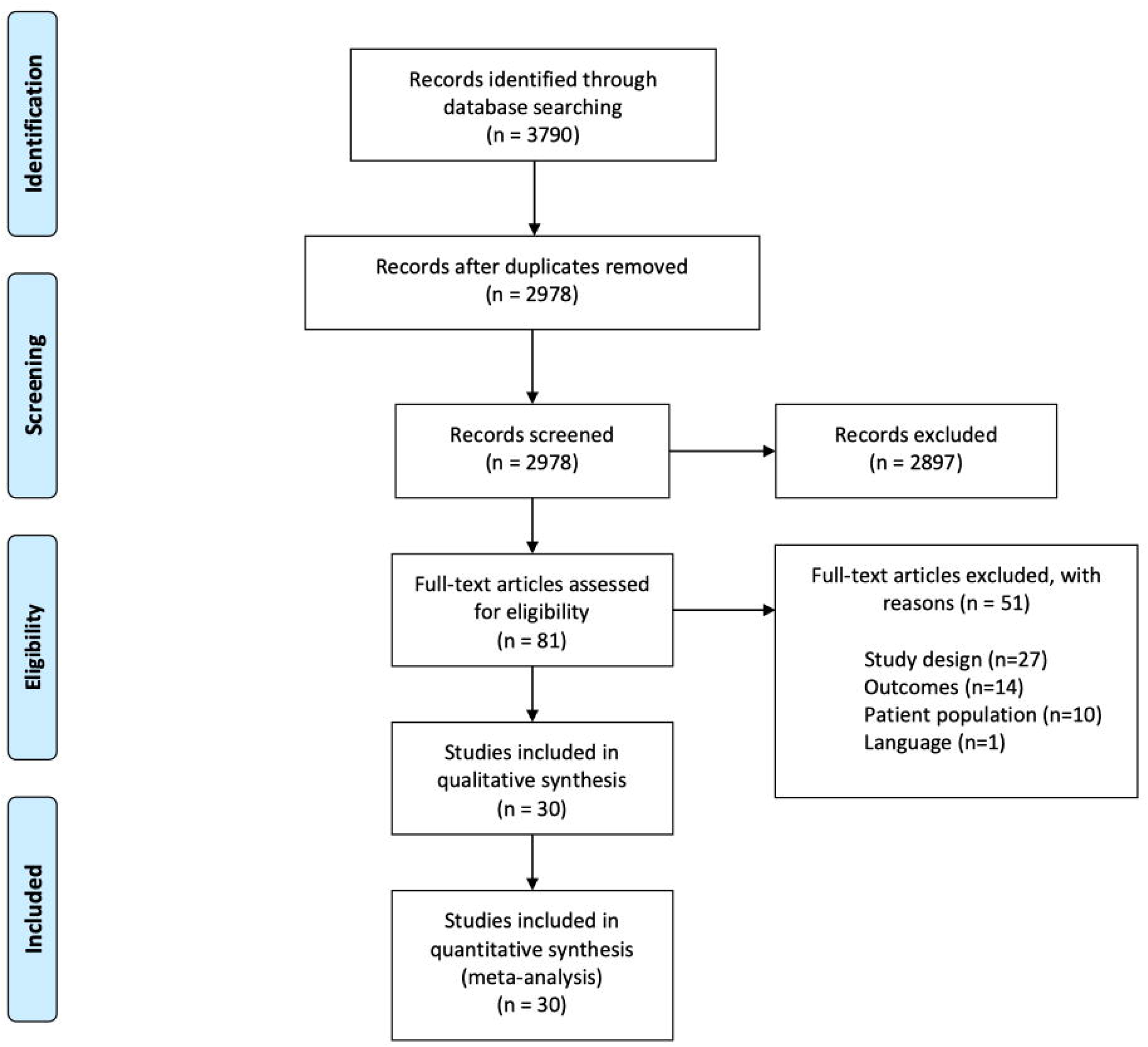

### Characteristics of included studies

Of the 30 studies, eight categories of study design were identified according to JBI methodological classification: retrospective cohort (n=8), retrospective case series (n=9), prospective cohort (n=7), cross-sectional (n=2), prospective case series (n=1), systematic review (n=1), meta-analysis (n=1), diagnostic and test accuracy (n=1). The median sample size and median study length with first and third quartile ranges were reported for each methodological category (Table 1). Studies originated from 13 different countries (Supplement 2. Table 1). According to World Bank Indexing, eight hold high-income status, two hold upper-middle-income status, one holds lower-middle-income status, and one (Taiwan) would hold high-income status if recognized by the United Nations as an independent member state. Two studies were published between 2005-2009, seven studies were published between 2010-2014, and 21 were published between 2015-2020 (Figure 2). Studies were published in 16 different journals, 30% (n=10) of which were published in the *Journal of Trauma and Acute Care Surgery*. The median impact factor (IF) was 3.3 with first and third interquartile ranges of 1.8 and 4.2, respectively (Supplement 2. Table 2).

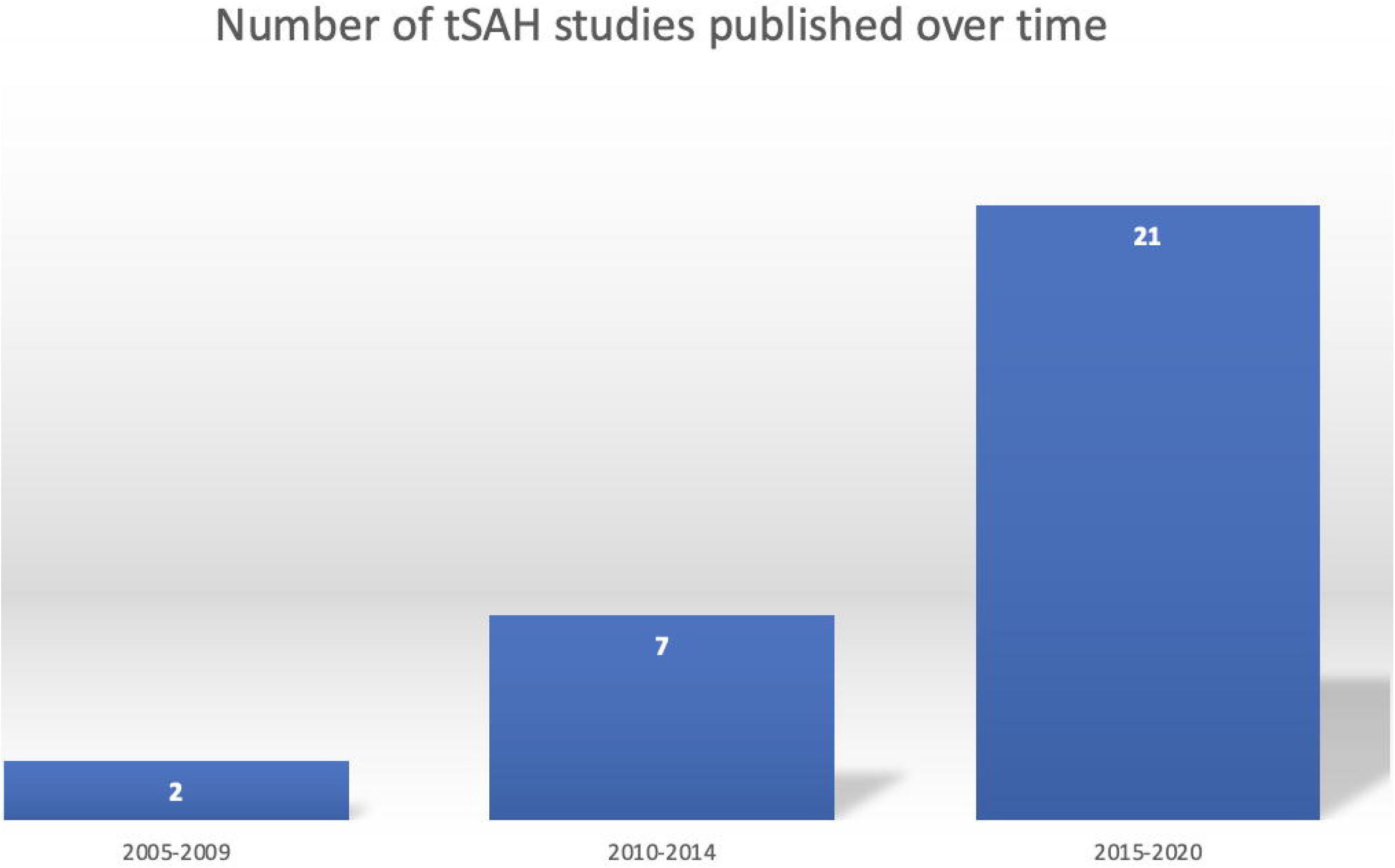

**Table 1.** Study Characteristics

| Study Type | Count | Median Sample Size (IQR) | Median Study Length (months, IQR) |
| --- | --- | --- | --- |
| Retrospective Case Series | 9 | 186 (63-473) | 60 (36-95) |
| Retrospective Cohort | 8 | 680 (452-1,103) | 68 (51-93) |
| Prospective Cohort | 7 | 161 (129-466) | 36 (36-84) |
| Cross-sectional | 2 | 1130 (767-1493) | 71 (47-95) |
| Prospective Case Series | 1 | 7 | 2 |
| Meta-analysis | 1 | 15,372 | N/A |
| Systematic Review | 1 | 1,074 | N/A |
| Diagnostic and Test Accuracy | 1 | 20 | 7 |

Studies were grouped into five separate categories: mild TBI (n=14), (Table 2); severe TBI (n=3), (Table 3); clinical management and prognosis (n=9), (Table 4); imaging (n=3), (Table 5); and traumatic aneurysm (n=1), (Table 6). All extraction data can be found in Supplement 2. Table 3.

**Table 2.**
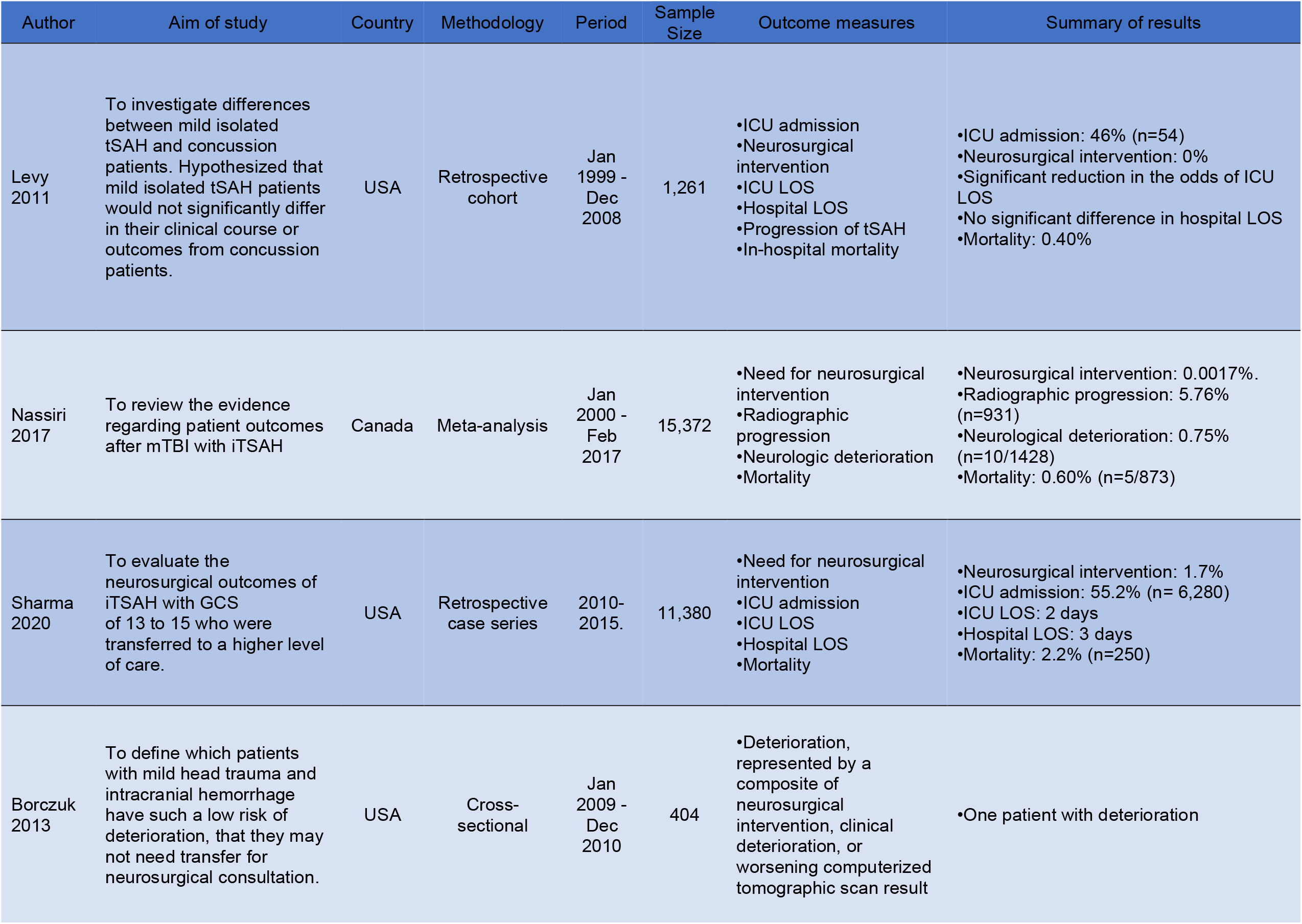
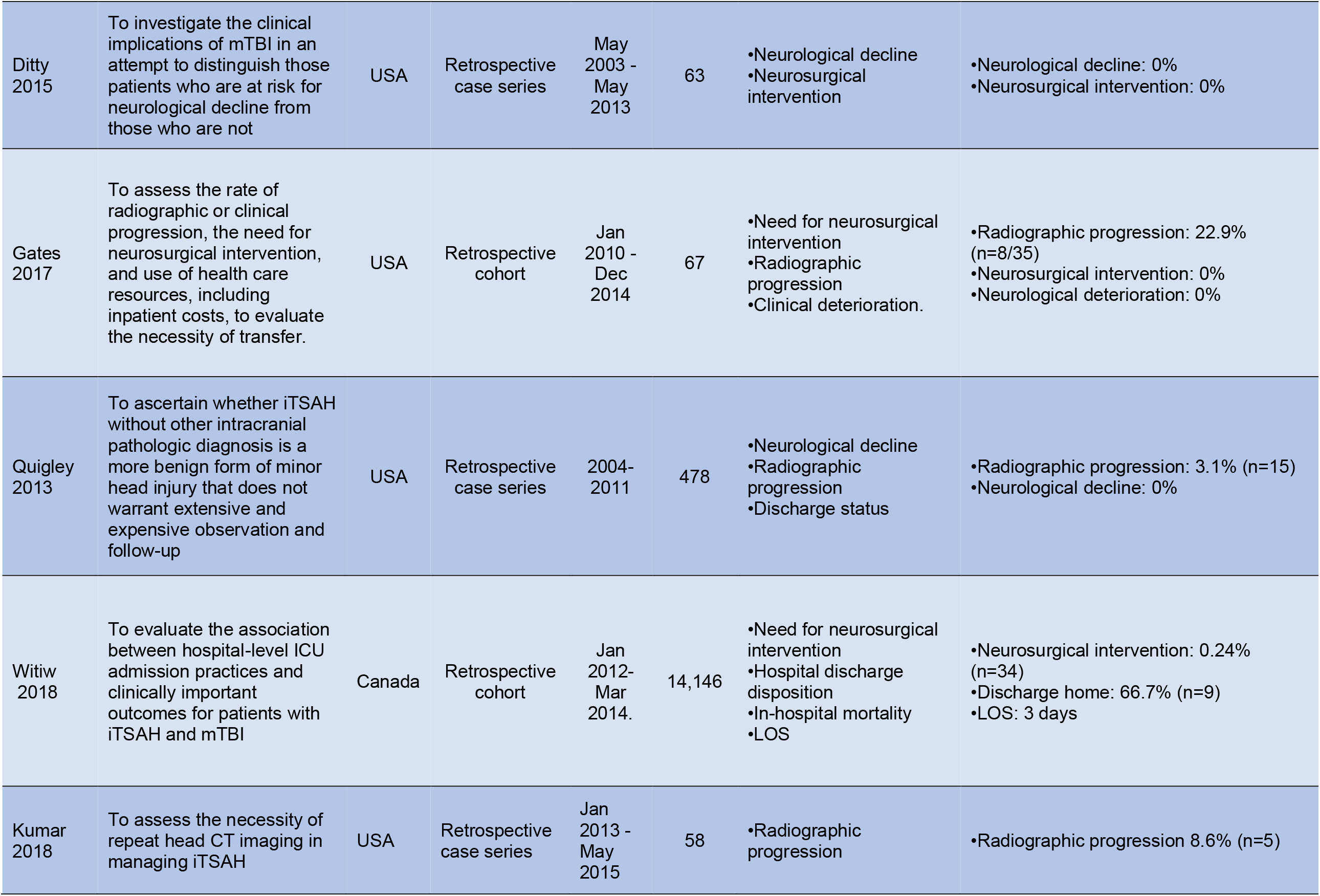
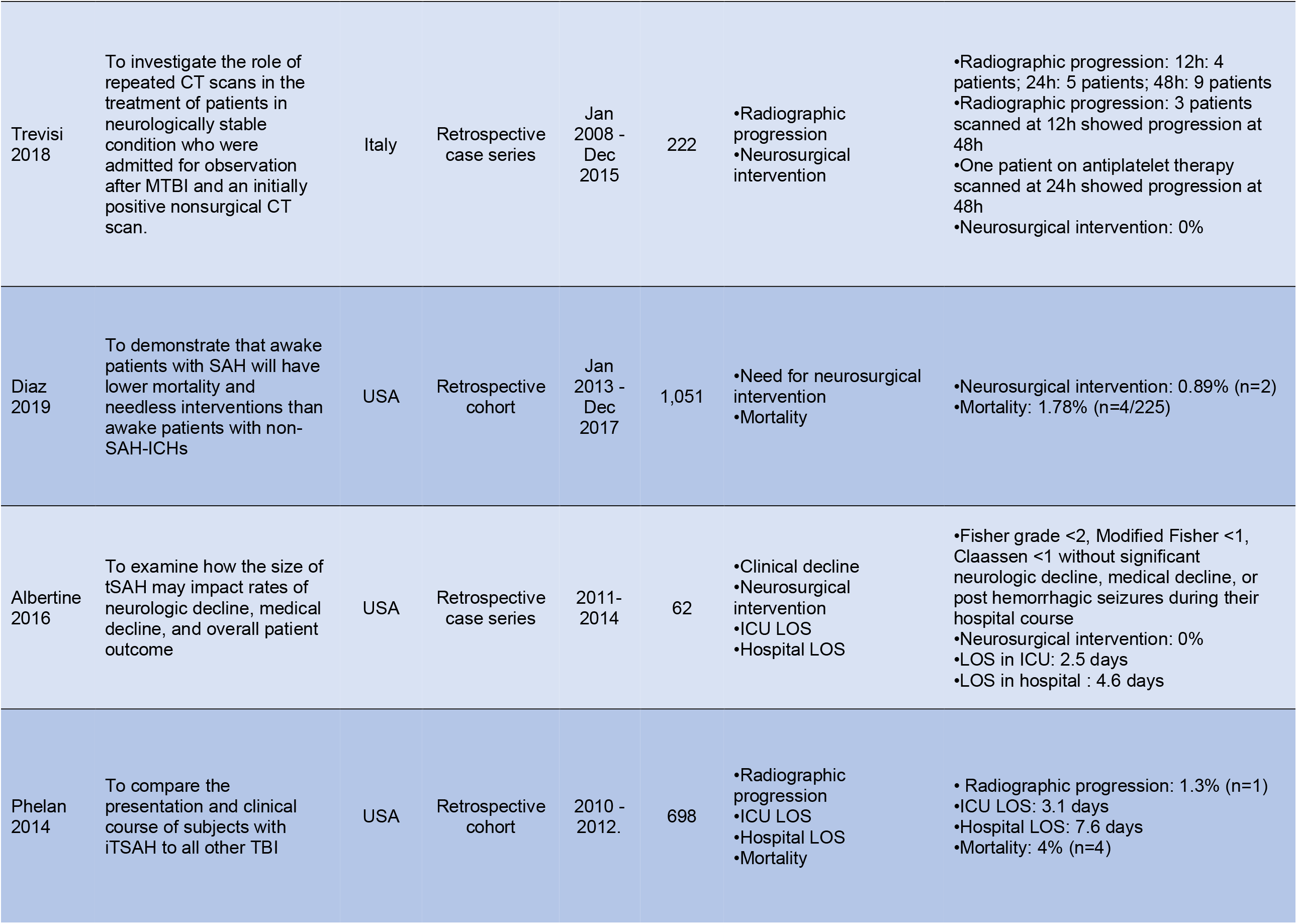
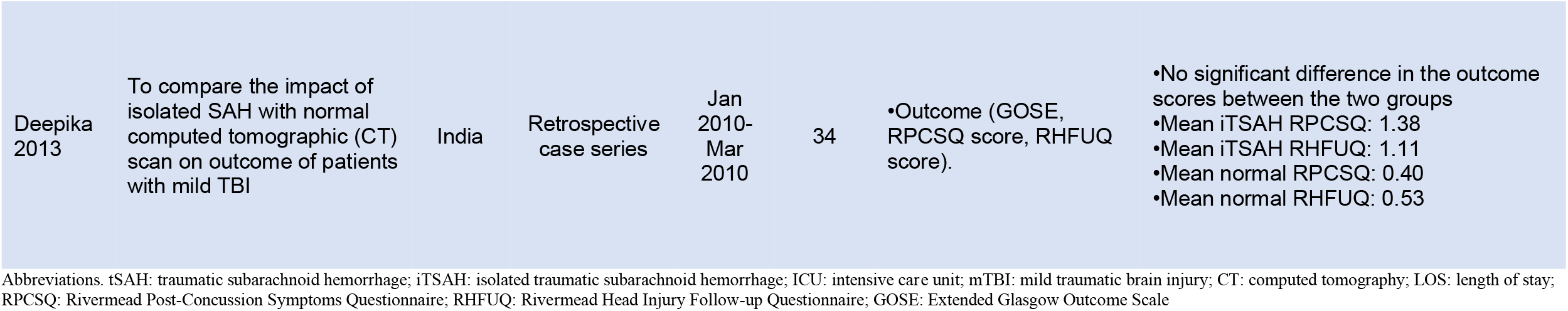
Mild TBI

| Author | Aim of study | Country | Methodology | Period | Sample Size | Outcome measures | Summary of results |
| --- | --- | --- | --- | --- | --- | --- | --- |
| Levy 2011 | To investigate differences between mild isolated tSAH and concussion patients. Hypothesized that mild isolated tSAH patients would not significantly differ in their clinical course or outcomes from concussion patients. | USA | Retrospective cohort | Jan 1999 - Dec 2008 | 1,261 | •ICU admission<br>•Neurosurgical intervention<br>•ICU LOS<br>•Hospital LOS<br>•Progression of tSAH<br>•In-hospital mortality | •ICU admission: 46% (n=54)<br>•Neurosurgical intervention: 0%<br>•Significant reduction in the odds of ICU LOS<br>•No significant difference in hospital LOS<br>•Mortality: 0.40% |
| Nassiri 2017 | To review the evidence regarding patient outcomes after mTBI with iTSAH | Canada | Meta-analysis | Jan 2000 - Feb 2017 | 15,372 | •Need for neurosurgical intervention<br>•Radiographic progression<br>•Neurologic deterioration<br>•Mortality | •Neurosurgical intervention: 0.0017%.<br>•Radiographic progression: 5.76% (n=931)<br>•Neurological deterioration: 0.75% (n=10/1428)<br>•Mortality: 0.60% (n=5/873) |
| Sharma 2020 | To evaluate the neurosurgical outcomes of iTSAH with GCS of 13 to 15 who were transferred to a higher level of care. | USA | Retrospective case series | 2010-2015. | 11,380 | •Need for neurosurgical intervention<br>•ICU admission<br>•ICU LOS<br>•Hospital LOS<br>•Mortality | •Neurosurgical intervention: 1.7%<br>•ICU admission: 55.2% (n= 6,280)<br>•ICU LOS: 2 days<br>•Hospital LOS: 3 days<br>•Mortality: 2.2% (n=250) |
| Borczuk 2013 | To define which patients with mild head trauma and intracranial hemorrhage have such a low risk of deterioration, that they may not need transfer for neurosurgical consultation. | USA | Cross-sectional | Jan 2009 - Dec 2010 | 404 | •Deterioration, represented by a composite of neurosurgical intervention, clinical deterioration, or worsening computerized tomographic scan result | •One patient with deterioration |
| Ditty<br>2015 | To investigate the clinical implications of mTBI in an attempt to distinguish those patients who are at risk for neurological decline from those who are not | USA | Retrospective case series | May 2003 - May 2013 | 63 | •Neurological decline<br>•Neurosurgical intervention | •Neurological decline: 0%<br>•Neurosurgical intervention: 0% |
| Gates<br>2017 | To assess the rate of radiographic or clinical progression, the need for neurosurgical intervention, and use of health care resources, including inpatient costs, to evaluate the necessity of transfer. | USA | Retrospective cohort | Jan 2010 - Dec 2014 | 67 | •Need for neurosurgical intervention<br>•Radiographic progression<br>•Clinical deterioration. | •Radiographic progression: 22.9% (n=8/35)<br>•Neurosurgical intervention: 0%<br>•Neurological deterioration: 0% |
| Quigley<br>2013 | To ascertain whether iTSAH without other intracranial pathologic diagnosis is a more benign form of minor head injury that does not warrant extensive and expensive observation and follow-up | USA | Retrospective case series | 2004-2011 | 478 | •Neurological decline<br>•Radiographic progression<br>•Discharge status | •Radiographic progression: 3.1% (n=15)<br>•Neurological decline: 0% |
| Witiw<br>2018 | To evaluate the association between hospital-level ICU admission practices and clinically important outcomes for patients with iTSAH and mTBI | Canada | Retrospective cohort | Jan 2012- Mar 2014. | 14,146 | •Need for neurosurgical intervention<br>•Hospital discharge disposition<br>•In-hospital mortality<br>•LOS | •Neurosurgical intervention: 0.24% (n=34)<br>•Discharge home: 66.7% (n=9)<br>•LOS: 3 days |
| Kumar<br>2018 | To assess the necessity of repeat head CT imaging in managing iTSAH | USA | Retrospective case series | Jan 2013 - May 2015 | 58 | •Radiographic progression | •Radiographic progression 8.6% (n=5) |
| Trevisi<br>2018 | To investigate the role of repeated CT scans in the treatment of patients in neurologically stable condition who were admitted for observation after MTBI and an initially positive nonsurgical CT scan. | Italy | Retrospective case series | Jan 2008 - Dec 2015 | 222 | •Radiographic progression<br>•Neurosurgical intervention | •Radiographic progression: 12h: 4 patients; 24h: 5 patients; 48h: 9 patients<br>•Radiographic progression: 3 patients scanned at 12h showed progression at 48h<br>•One patient on antiplatelet therapy scanned at 24h showed progression at 48h<br>•Neurosurgical intervention: 0% |
| Diaz<br>2019 | To demonstrate that awake patients with SAH will have lower mortality and needless interventions than awake patients with non-SAH-ICHs | USA | Retrospective cohort | Jan 2013 - Dec 2017 | 1,051 | •Need for neurosurgical intervention<br>•Mortality | •Neurosurgical intervention: 0.89% (n=2)<br>•Mortality: 1.78% (n=4/225) |
| Albertine<br>2016 | To examine how the size of tSAH may impact rates of neurologic decline, medical decline, and overall patient outcome | USA | Retrospective case series | 2011-2014 | 62 | •Clinical decline<br>•Neurosurgical intervention<br>•ICU LOS<br>•Hospital LOS | •Fisher grade <2, Modified Fisher <1, Claassen <1 without significant neurologic decline, medical decline, or post hemorrhagic seizures during their hospital course<br>•Neurosurgical intervention: 0%<br>•LOS in ICU: 2.5 days<br>•LOS in hospital : 4.6 days |
| Phelan<br>2014 | To compare the presentation and clinical course of subjects with ITSAH to all other TBI | USA | Retrospective cohort | 2010 - 2012. | 698 | •Radiographic progression<br>•ICU LOS<br>•Hospital LOS<br>•Mortality | • Radiographic progression: 1.3% (n=1)<br>•ICU LOS: 3.1 days<br>•Hospital LOS: 7.6 days<br>•Mortality: 4% (n=4) |
| Deepika<br>2013 | To compare the impact of isolated SAH with normal computed tomographic (CT) scan on outcome of patients with mild TBI | India | Retrospective case series | Jan 2010-Mar 2010 | 34 | •Outcome (GOSE, RPCSQ score, RHFUQ score). | •No significant difference in the outcome scores between the two groups<br>•Mean iTSAH RPCSQ: 1.38<br>•Mean iTSAH RHFUQ: 1.11<br>•Mean normal RPCSQ: 0.40<br>•Mean normal RHFUQ: 0.53 |

Abbreviations. tSAH: traumatic subarachnoid hemorrhage; iTSAH: isolated traumatic subarachnoid hemorrhage; ICU: intensive care unit; mTBI: mild traumatic brain injury; CT: computed tomography; LOS: length of stay; RPCSQ: Rivermead Post-Concussion Symptoms Questionnaire; RHFUQ: Rivermead Head Injury Follow-up Questionnaire; GOSE: Extended Glasgow Outcome Scale

**Table 3.** Severe TBI

| Author | Aim of study | Country | Methodology | Period | Sample Size | Outcome measures | Summary of findings |
| --- | --- | --- | --- | --- | --- | --- | --- |
| Thelin 2017 | To evaluate the Stockholm and Helsinki CT scores for predicting functional outcome, in comparison with the Rotterdam CT score and Marshall CT classification. To assess which individual components of the CT scores best predict outcome and what additional prognostic value the CT scoring systems contribute to a clinical prognostic model. | United Kingdom | Prospective cohort | 2005-2014 | 1,115 | •CT severity score as predictor of GOS | •tSAH was the most important component of the Stockholm and Helsinki CT scoring system for outcome prediction |
| Matsushima 2015 | To investigate the relationship between time to surgery and outcomes in patients with isolated sTBI requiring an emergent neurosurgical intervention. | USA | Prospective cohort | 2003 - 2013 | 161 | •In-hospital mortality | •In-hospital mortality rate: 34.5% (early) vs. 59.1% (late)<br>•Neurosurgical intervention was significantly associated with a higher odds of patient survival |
| Tu 2011 | To evaluate whether the maximum thickness of subarachnoid blood is an independent prognostic marker of mortality after tSAH | China | Prospective cohort | 2007-2010 | 104 | •1-month mortality | •Maximum thickness of subarachnoid blood is a prognostic marker of 1-month mortality |

Abbreviations. tSAH: traumatic subarachnoid hemorrhage; CT: computed tomography

**Table 4.**
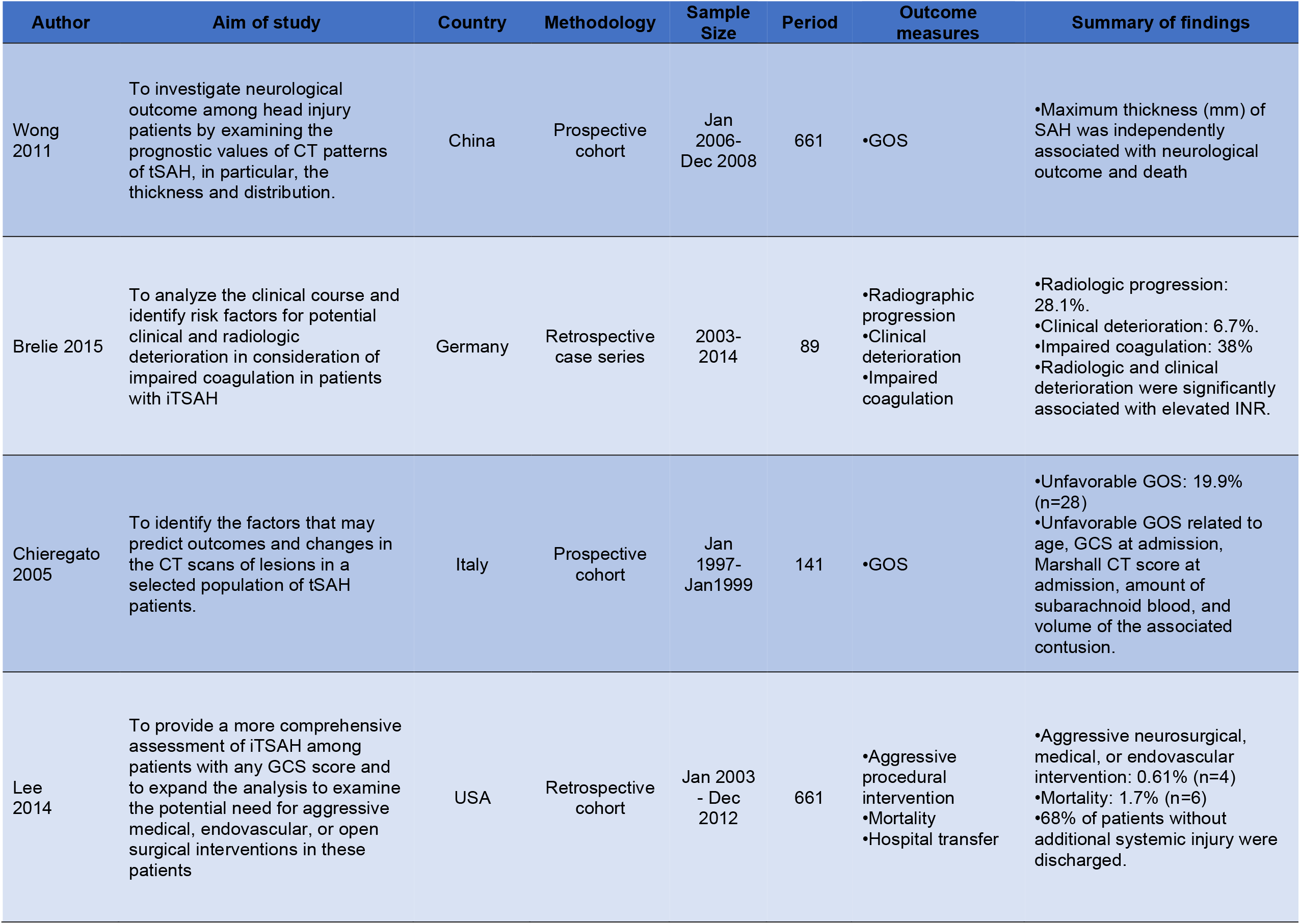
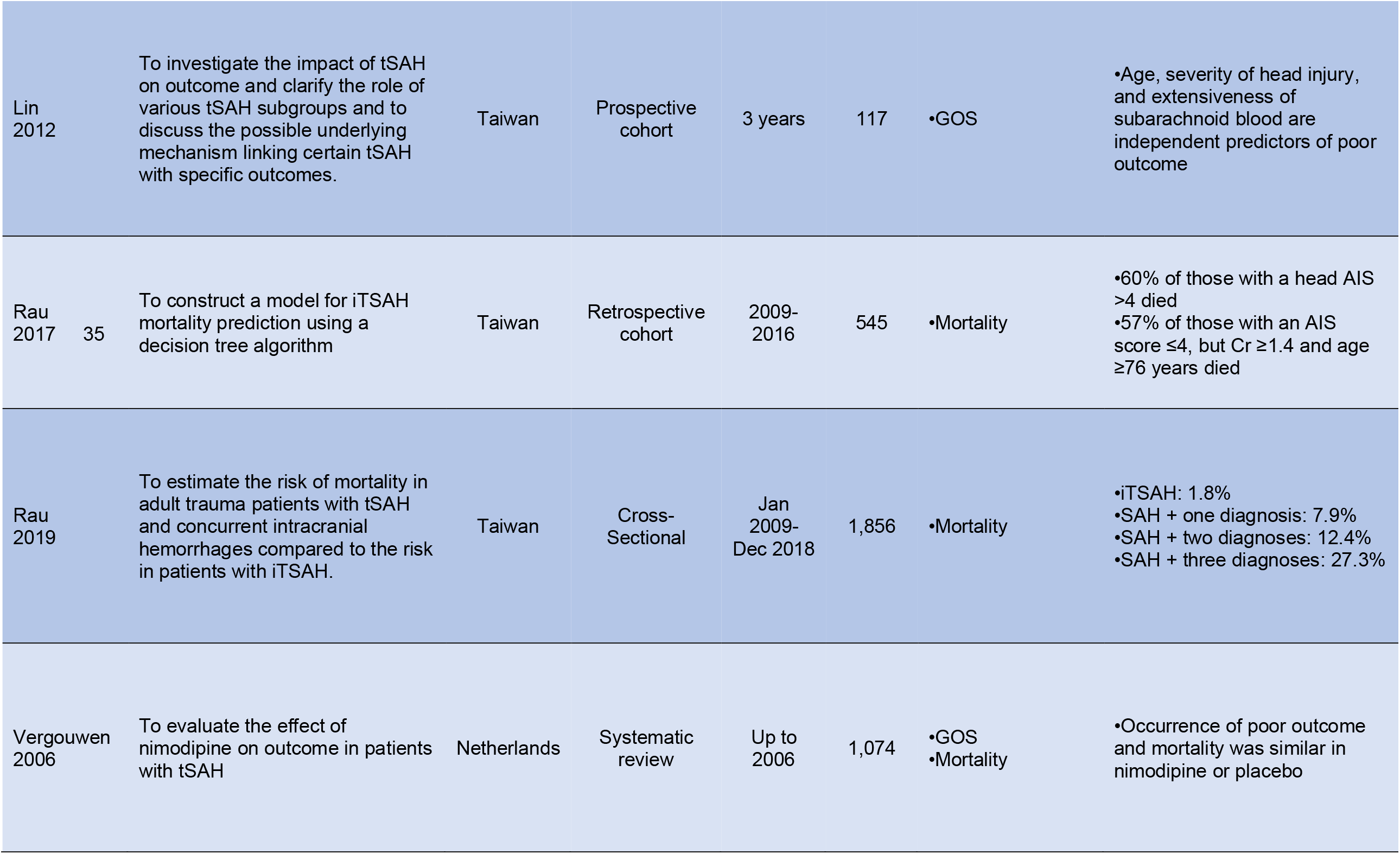
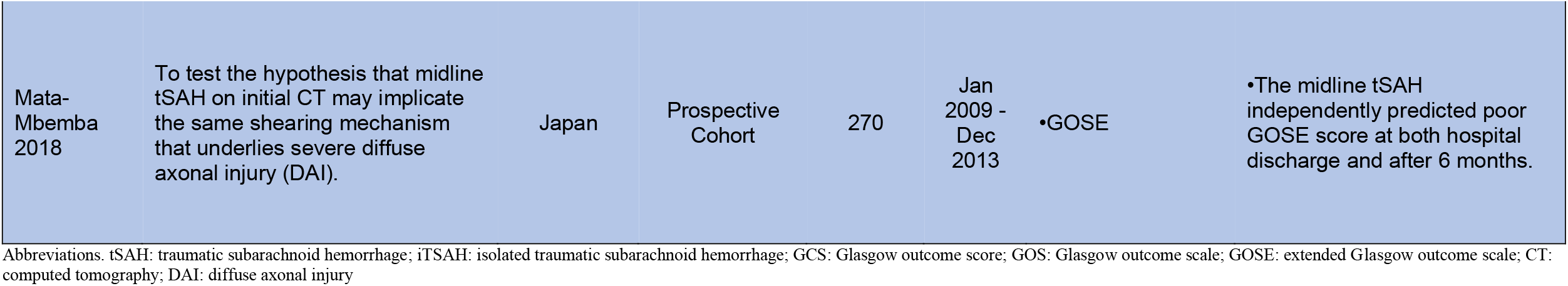
Clinical Management and Diagnosis

| Author | Aim of study | Country | Methodology | Sample Size | Period | Outcome measures | Summary of findings |
| --- | --- | --- | --- | --- | --- | --- | --- |
| Wong 2011 | To investigate neurological outcome among head injury patients by examining the prognostic values of CT patterns of tSAH, in particular, the thickness and distribution. | China | Prospective cohort | Jan 2006- Dec 2008 | 661 | •GOS | •Maximum thickness (mm) of SAH was independently associated with neurological outcome and death |
| Brelie 2015 | To analyze the clinical course and identify risk factors for potential clinical and radiologic deterioration in consideration of impaired coagulation in patients with iTSAH | Germany | Retrospective case series | 2003-2014 | 89 | •Radiographic progression<br>•Clinical deterioration<br>•Impaired coagulation | •Radiologic progression: 28.1%.<br>•Clinical deterioration: 6.7%.<br>•Impaired coagulation: 38%<br>•Radiologic and clinical deterioration were significantly associated with elevated INR. |
| Chierigato 2005 | To identify the factors that may predict outcomes and changes in the CT scans of lesions in a selected population of tSAH patients. | Italy | Prospective cohort | Jan 1997- Jan1999 | 141 | •GOS | •Unfavorable GOS: 19.9% (n=28)<br>•Unfavorable GOS related to age, GCS at admission, Marshall CT score at admission, amount of subarachnoid blood, and volume of the associated contusion. |
| Lee 2014 | To provide a more comprehensive assessment of iTSAH among patients with any GCS score and to expand the analysis to examine the potential need for aggressive medical, endovascular, or open surgical interventions in these patients | USA | Retrospective cohort | Jan 2003 - Dec 2012 | 661 | •Aggressive procedural intervention<br>•Mortality<br>•Hospital transfer | •Aggressive neurosurgical, medical, or endovascular intervention: 0.61% (n=4)<br>•Mortality: 1.7% (n=6)<br>•68% of patients without additional systemic injury were discharged. |

|  |  |  |  |  |  |  |  |  |
| --- | --- | --- | --- | --- | --- | --- | --- | --- |
| Lin<br>2012 |  | To investigate the impact of tSAH on outcome and clarify the role of various tSAH subgroups and to discuss the possible underlying mechanism linking certain tSAH with specific outcomes. | Taiwan | Prospective cohort | 3 years | 117 | •GOS | •Age, severity of head injury, and extensiveness of subarachnoid blood are independent predictors of poor outcome |
| Rau<br>2017 | 35 | To construct a model for iTSAH mortality prediction using a decision tree algorithm | Taiwan | Retrospective cohort | 2009-2016 | 545 | •Mortality | •60% of those with a head AIS >4 died<br>•57% of those with an AIS score ≤4, but Cr ≥1.4 and age ≥76 years died |
| Rau<br>2019 |  | To estimate the risk of mortality in adult trauma patients with tSAH and concurrent intracranial hemorrhages compared to the risk in patients with iTSAH. | Taiwan | Cross-Sectional | Jan 2009-Dec 2018 | 1,856 | •Mortality | •iTSAH: 1.8%<br>•SAH + one diagnosis: 7.9%<br>•SAH + two diagnoses: 12.4%<br>•SAH + three diagnoses: 27.3% |
| Vergouwen<br>2006 |  | To evaluate the effect of nimodipine on outcome in patients with tSAH | Netherlands | Systematic review | Up to 2006 | 1,074 | •GOS<br>•Mortality | •Occurrence of poor outcome and mortality was similar in nimodipine or placebo |
| Mata-Mbemba 2018 | To test the hypothesis that midline tSAH on initial CT may implicate the same shearing mechanism that underlies severe diffuse axonal injury (DAI). | Japan | Prospective Cohort | 270 | Jan 2009 - Dec 2013 | •GOSE | •The midline tSAH independently predicted poor GOSE score at both hospital discharge and after 6 months. |

Abbreviations. tSAH: traumatic subarachnoid hemorrhage; iSAH: isolated traumatic subarachnoid hemorrhage; GCS: Glasgow outcome score; GOS: Glasgow outcome scale; GOSE: extended Glasgow outcome scale; CT: computed tomography; DAI: diffuse axonal injury

**Table 5.**
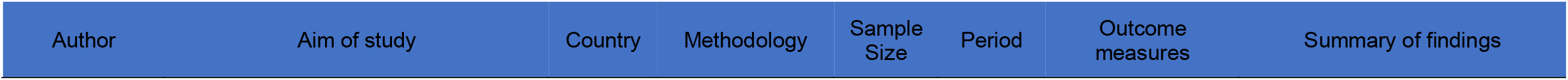
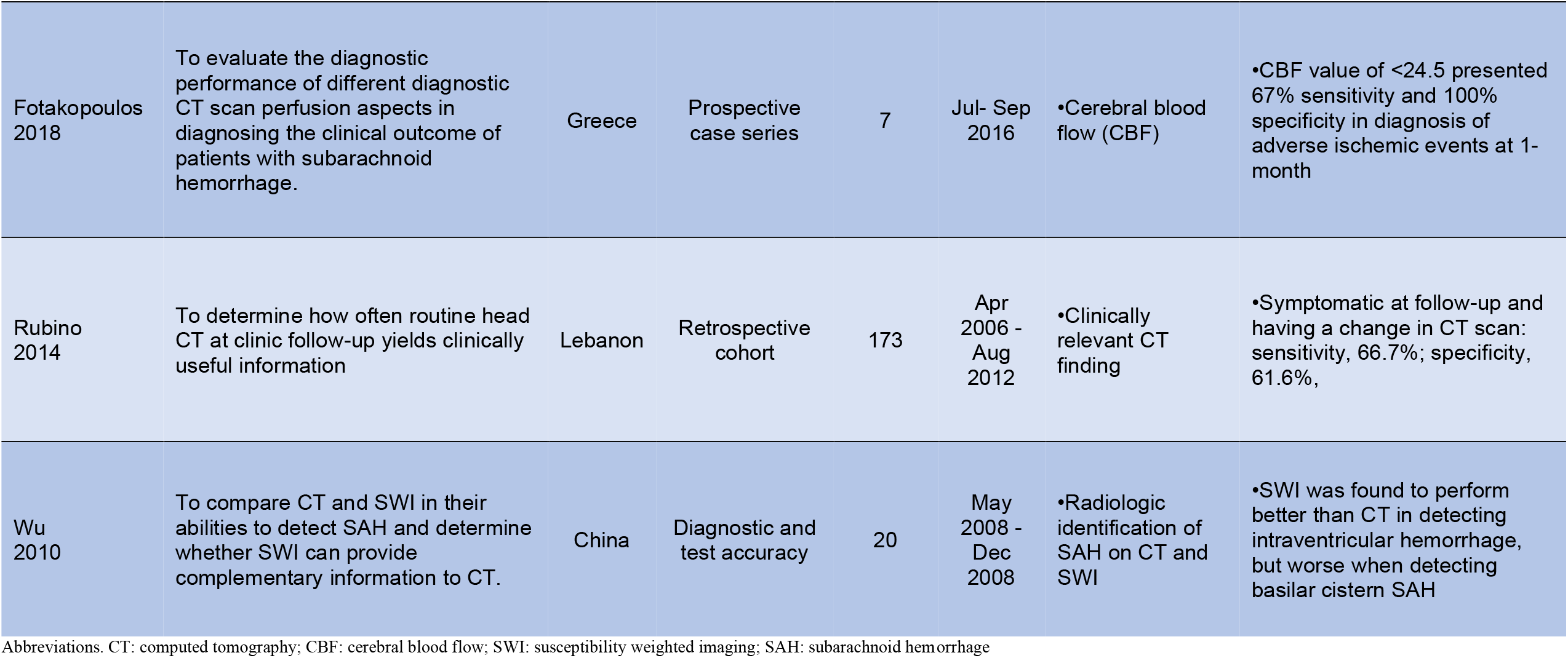
Imaging

| Author | Aim of study | Country | Methodology | Sample Size | Period | Outcome measures | Summary of findings |
| --- | --- | --- | --- | --- | --- | --- | --- |
| Fotakopoulos<br>2018 | To evaluate the diagnostic performance of different diagnostic CT scan perfusion aspects in diagnosing the clinical outcome of patients with subarachnoid hemorrhage. | Greece | Prospective case series | 7 | Jul- Sep 2016 | •Cerebral blood flow (CBF) | •CBF value of <24.5 presented 67% sensitivity and 100% specificity in diagnosis of adverse ischemic events at 1-month |
| Rubino<br>2014 | To determine how often routine head CT at clinic follow-up yields clinically useful information | Lebanon | Retrospective cohort | 173 | Apr 2006 - Aug 2012 | •Clinically relevant CT finding | •Symptomatic at follow-up and having a change in CT scan: sensitivity, 66.7%; specificity, 61.6%, |
| Wu<br>2010 | To compare CT and SWI in their abilities to detect SAH and determine whether SWI can provide complementary information to CT. | China | Diagnostic and test accuracy | 20 | May 2008 - Dec 2008 | •Radiologic identification of SAH on CT and SWI | •SWI was found to perform better than CT in detecting intraventricular hemorrhage, but worse when detecting basilar cistern SAH |

Abbreviations. CT: computed tomography; CBF: cerebral blood flow; SWI: susceptibility weighted imaging; SAH: subarachnoid hemorrhage

**Table 6.**
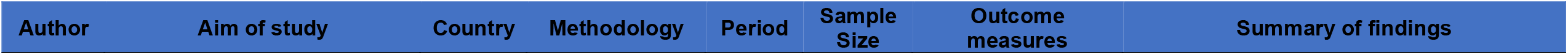
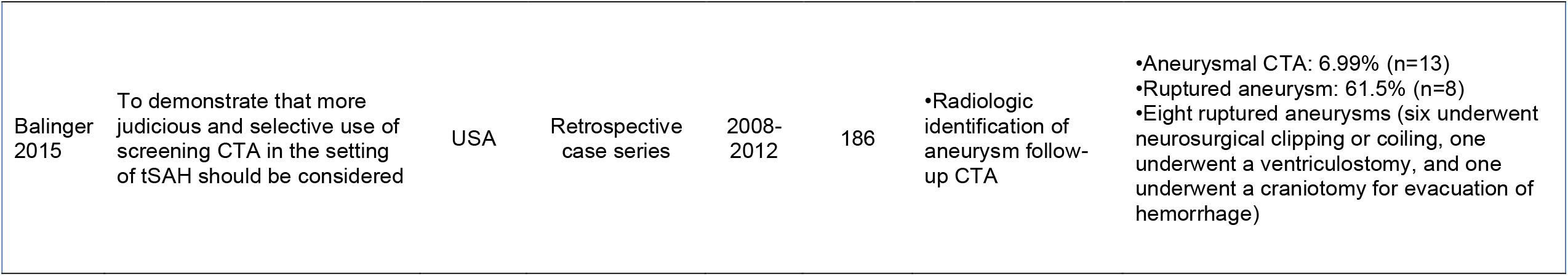
Aneurysmal tSAH

### Review findings

#### Mild TBI

Studies were categorized as mild TBI if the GCS of the patient population was 13-15 on presentation (Table X).

A meta-analysis of 15,372 tSAH patients found the incidence of need for eventual neurosurgical intervention to be 0.0017%.^12^ They also found a 5.76% (n=931) incidence for radiographic progression, 0.75% (n=10/1428) incidence for neurological deterioration, and 0.60% (n=5/873) incidence for mortality.

A retrospective cohort study of 14,146 patients with mTBI and iTSAH found the need for neurosurgical intervention to be 0.24% (n=34).^13^ The discharge disposition was 66.7% (n=9) to home under self-care and 8.8% (n=1) to home with support. Median hospital LOS for all subjects was three days (IQR, 2–5 d).

A retrospective cohort study of 102 iTSAH patients, of which 77 had a mTBI, found that 27 underwent a routine repeat CT scan.^9^ The radiographic progression rate was 11% (n=3). The other 50 patients were observed until a change occurred in their neurological exam. 8% (n=4) of these patients needed a repeat CT scan and 2% (n=1) demonstrated radiographic progression. A total of 1.3% (n=1) had clinical or radiographic hemorrhage progression. The ICU LOS was 3.1, hospital LOS was 7.6, and 4% (n=4) of the patients died.

A retrospective cohort study comparing 225 iTSAH patients and 826 non-SAH-ICH patients found that 0.89% (n=2) tSAH patients and 12.1% (n=100) of non-SAH-ICH patients required neurosurgical intervention.^14^ Mortality rates were 1.78% (n=4/225) in the SAH patients and 2.66% (n=22/826) in the non-SAH-ICH patients.

A retrospective cohort study of 117 iTSAH patients and 1144 patients with concussion found that 46% (n=54) of iTSAH patients and 7% (n=83) of concussion patients were admitted to the ICU.^15^ No patients required neurosurgical intervention.

A retrospective cohort study of 67 iTSAH patients found none to require neurosurgical intervention.^16^ Radiographic progression was observed in eight of the 35 patients who were rescanned. The mean age in those patients was 74 years compared to 62 years in those who were not rescanned. Six of the eight patients had a slight expansion in the subarachnoid space or new intraventricular blood. One patient experienced neurological deterioration.

A retrospective cohort study of 41 iTSAH patients found none to require neurosurgical intervention.^17^ Nor did any patients experience neurological worsening.

A cross-sectional study of 75 iTSAH patients found that one 98-year-old patient experienced neurological deterioration (OR, 0.08) but did not require neurosurgical intervention.^8^

A retrospective case series of 11,380 iTSAH patients requiring transfer to a higher-level facility found that 1.7% of patients required neurosurgical intervention.^18^ 55.2% (n= 6,280) of the patients were admitted to the ICU, where their LOS was a median of two days. 62.2 % (n=7,077) of the patients were discharged home without any services. 2.2% (n=250) of the patients died.

A retrospective case series of 478 iTSAH patients found that 98.3% (n=470) had a second post-admission CT scan.^19^ 3.1% (n=15) had CT progression. No patients experienced neurological decline nor required neurosurgical intervention.

A retrospective case series of 109 iTSAH patients found that at 12-hours post first scan, radiographic progression was found in four patients.^20^ Radiographic progression was seen in five patients 24-hours post first scan, and in nine patients at 48-hours post first scan. Three patients scanned at 12-hours showed progression at 48-hours. One patient on antiplatelet therapy scanned at 24-hours showed progression at 48-hours. No patient required neurosurgical intervention.

A retrospective case series of 58 iTSAH patients found that radiographic progression was seen in 8.6% (n=5) of patients. These patients did not require re-admission. No patients required neurosurgical intervention.^21^

A retrospective case series of 62 iTSAH patients found that patients with lower-grade tSAHs (Fisher grade <2, Modified Fisher <1, Claassen <1) did not show significant neurologic decline, medical decline, or post-hemorrhagic seizures during their hospital course.^22^ Median ICU LOS was 2.5 days. No patients required neurosurgical intervention.

A retrospective case series of 34 iTSAH patients found that compared with mTBI patients with normal CT scans, there was no significant difference in the outcome scores between the two groups one-year after injury.^23^ The telephonic Glasgow Outcome Scale-Extended (GOSE), Rivermead Head Injury Follow-up Questionnaire (RHFUQ), and Rivermead Post-Concussion Symptoms Questionnaire (RPCSQ) scores were used to assess outcome. The mean RHFUQ and RPCSQ scores for patients with isolated SAH were 1.11 and 1.38, respectively. The mean RHFUQ and RPCSQ scores for patients with normal CT scans were 0.53 and 0.40, respectively.

#### Severe TBI

Studies were categorized as severe TBI if the GCS of the patient population was <8 on presentation.

A prospective cohort study of 1,115 sTBI patients found that tSAH was the most essential component of the Stockholm and Helsinki CT scoring system for outcome prediction. The aggregate SAH component of the Stockholm CT score was the strongest predictor of unfavorable outcome.^24^

A prospective cohort study of 161 sTBI patients found 55.5% to have iTSAH on CT imaging. In-hospital mortality of patients whose surgery lasted less than 200 minutes was compared with those whose surgery lasted 200 minutes or longer. The in-hospital mortality rate was significantly lower in the early group (34.5% vs. 59.1%, p = 0.03). Early neurosurgical intervention was significantly associated with higher odds of patient survival (odds ratio, 7.41, p = 0.009).^25^

A prospective cohort study of 125 iTSAH patients found that the maximum thickness of subarachnoid blood is an independent prognostic marker of 1-month mortality after traumatic subarachnoid hemorrhage.^26^

#### Imaging

A retrospective cohort study of 75 iTSAH patients with at least one clinic follow-up found that the sensitivity and specificity of being symptomatic at follow-up and having a CT scan change was 66.7% and 61.6%, respectively.^27^

A diagnostic and test accuracy study of 20 tSAH patients found that Susceptibility-Weighted Imaging (SWI) is very sensitive to small amounts of SAH and is useful in differentiating SAH from veins.^28^ SWI performed better than CT in detecting intraventricular hemorrhage but worse when detecting basilar cistern SAH.

A prospective case series of 7 tSAH patients found that cerebral blood flow (CBF), as derived from CT perfusion, is a measurable index that may help detect the degree of very early cerebral ischemia in patients suffering from SAH.^29^ A CBF value of <24.5 presented 67% sensitivity and 100% specificity in diagnosing adverse ischemic events at 1-month (p=0.041).

#### Clinical management and prognosis

A systematic review of 1,074 tSAH patients treated with nimodipine or placebo found that the occurrence of poor outcome and mortality was similar in both groups.^30^

A prospective cohort study of 661 tSAH patients found that the maximum thickness (mm) of SAH was independently associated with neurological outcome (OR 0.8) and death (OR 1.3), but not with the extent or location of hemorrhage.^31^

A prospective cohort study of 270 tSAH patients who underwent brain MRI within 30 days found that 28.5% (n=77) had diffuse axonal injury (DAI) and that tSAH was independently associated with DAI.^32^ Midline tSAH was independently associated with both overall DAI and DAI stage two or three. The midline tSAH on initial CT had a sensitivity of 60.8%, a specificity of 81.7%, and positive and negative predictive values of 43.7% and 89.9%, respectively, for severe DAI. When adjusted for admission GCS, the midline tSAH independently predicted low GOSE score at both hospital discharge and after six months.

A prospective cohort study of 141 tSAH patients found that unfavorable GOS was significantly related to age, GCS at admission, Marshall CT score at admission, amount of subarachnoid blood, and volume of the associated contusion.^33^

A prospective cohort study of 117 tSAH patients found that age, severity of head injury, and subarachnoid blood extensiveness are independent predictors of poor outcome.^34^ Patients with extensive tSAH with intraventricular hemorrhage tend to be associated with vasospasm in the acute stage.

A retrospective cohort study of 661 iTSAH patients found that only 0.61% (n=4) patients underwent any sort of aggressive neurosurgical, medical, or endovascular intervention. 1.7% (n=6) of patients died in hospital, and five of the six were older than 80. 68% of patients without additional systemic injury were discharged, including 53% of patients with a GCS score 3-8. Age, severity of injury, extensive SAH, and IVH are independent predictors of poor outcome in the cohort of tSAH patients. Patients with extensive tSAH and intraventricular hemorrhage tend to be associated with vasospasm in the acute phase.^35^

A retrospective cohort study of 546 iTSAH patients found that of the patients with isolated tSAH, 60% of those with a head AIS >4 died. The mortality was 57% for patients with an AIS score ≤4 in addition to having a Cr ≥1.4 and age ≥76 years.^36^ All patients without the above criteria survived.

A retrospective case series of 89 iTSAH patients with GCS >8 and a follow-up CT scan found that the radiologic expansion or conversion rate of the SAH was 28.1%.^37^ The rate of clinical deterioration was 6.7%. Neither the initial pattern intracranial localization nor the number of sulci involved with the iTSAH was associated with clinical worsening. 38% of patients had impaired coagulation; 17.9% of patients had elevated INR. Radiologic and clinical deterioration was significantly associated with elevated INR.

A cross-sectional study of 1,856 tSAH patients found that patients with iTSAH had a 1.8% rate of mortality, compared with patients with SAH + one diagnosis (7.9%), SAH + two diagnoses (12.4%), and SAH + three diagnoses (27.3%), where one, two, and three diagnoses indicated the existance of one, two, or three other types of intracranial hemorrhage (SDH, EDH, or ICH).^38^ When controlling for sex, age, and pre-existing comorbidities, Group II, Group III, and Group IV patients had a 4.0, 8.9, and 21.1 times higher adjusted odds ratio for mortality, respectively, than the patients with iTSAH.

#### Traumatic aneurysm

A retrospective case series of 186 tSAH patients who underwent CT angiography (CTA) found 7% (n=13) of patients had an aneurysm on the follow-up CTA.^39^ Thirteen patients (6.99%) had an aneurysm on the follow-up CTA. Of those, 61.5% (n=8) presented with a ruptured aneurysm. All eight patients with a ruptured aneurysm had central SAH (CSH), whereas none of the five patients with an unruptured aneurysm had CSH. The mechanism of injury did not correlate with the presence of an aneurysm.

## Discussion

This scoping review sought to elucidate the available evidence regarding the diagnosis and management of tSAH. Ultimately, we found little evidence regarding diagnostic capability. Most studies focused on clinical prognosis, management, and outcomes. We found that the studies could be grouped into five categories: tSAH associated with mTBI; tSAH associated with sTBI; tSAH and imaging; clinical prognosis, management, and outcomes; and post-tSAH aneurysm. This discussion focuses on the evidence discovered for each of the five categories, the scoping review’s limitations, and recommendations for future research.

### Study Characteristics

TSAH is a topic that is rapidly increasing in interest. Only two studies on the topic were published between 2005 and 2009, with an increase to seven studies published between 2010 and 2014. From 2015 to 2020, that number has almost quadrupled, rising to 21 published studies. While this is notable, the majority of these studies come from high-income countries. Excluding China, which is set to become a high-income country within the next couple of years, only two studies are from an LMIC, Lebanon, an upper-middle-income country, and India, a lower-middle-income country. An estimated 90% of injury-related deaths occur in LMICs, and the resultant social and economic costs are highest for these communities.^40–42^ More research is needed to quantify the social and economic impact of the mortality rate of tSAH and its associated physical and mental disability.

It is important to note that among the 30 studies, there were only two systematic reviews, one of which was a meta-analysis. The retrospective nature of most studies limits the strength of the evidence. Eight studies were retrospective case series, which inherently carries a high-level selection bias and no comparison group.

### Mild TBI

There were 14 studies whose patient population consisted of individuals presenting with GCS 13-15. Overall, patients from this population were found to have very low rates of need for neurosurgical intervention. One study found that trauma patients with a GCS score of 13–15 on initial evaluation and imaging evidence of traumatic SAH are doubtful to require neurosurgical consultation or transfer to tertiary care centers and can safely be discharged, barring the presence of other injuries or medical issues that require inpatient management.^17^

Multiple other studies either found patients in this group to not require or rarely require neurosurgical management.^8,12,13,18^ Patients from this population rarely experience neurological decline, need for transfer to a higher-level facility, or undergo radiographic progression. One study found no difference in the outcome of patients with isolated SAH compared with those with normal CT scans one-year after injury.^23^ One study found that the rate of neurological deterioration due to an expansion of iTSAH in patients with mTBI is low, regardless of antiplatelet and anticoagulant agents.^16^ Another study found that patients with low-grade tSAH did not demonstrate any neurological decline, medical decline, post-traumatic seizure, and most of these patients demonstrate good clinical outcomes. This suggests that patients may not require as aggressive monitoring as is currently provided for those with tSAH.^22^ CT imaging may have little efficacy in changing mild iTSAH management and is poorly correlated with clinical progression. A less aggressive management protocol may be more appropriate in managing these patients.^21^Another study found that no lesions showed significant worsening on repeat imaging at 24 or 48 hours post initial scan. Given neurologic stability, a control scan can be safely delayed as long as 48 hours to avoid an excessive number of unnecessary scans.^20^

Multiple studies reported high rates of admission to the ICU. One study showed that tSAH patients had increased odds of admission to the ICU, yet their ICU LOS was short. This suggests that healthcare facilities should consider creating ICU criteria for the mTBI population to optimize ICU utilization. Variables such as age, comorbidities, and neurologic condition could be more important indicators to consider rather than the presence of a small volume of blood in the subarachnoid space when admitting mTBI patients to the ICU.^15^ Clinical experience and acumen should be used to guide decision-making regarding ICU admission in this patient population. However, the presence of isolated tSAH should not, of itself, represent an indication to admit a patient with a clinically mild TBI to the critical care unit. Reevaluation of hospital-level practices may represent an opportunity for greater resource optimization.^13^

### Severe TBI

There is a paucity of information available for tSAH associated with sTBI. The findings were limited to three studies. One found a link between a shorter time-interval to surgical intervention and improved mortality.^25^ Most studies evaluating time to treatment and SAH involve aneurysmal SAH.^43–45^ This is particularly important for patients who present to the ED with low GCS and high-risk TBI mechanism so that the diagnostic and therapeutic processes include initial resuscitation, CT scan, neurosurgery consultation, and transfer the operating room can be expedited. A second study found that the Stockholm and Helsinki CT scores performed better than the Rotterdam and Marshall CT scores.^24^ It may be warranted to explore a move towards these new scoring systems to effectively target intensive care and surgical treatment as early as possible. The third study showed that the maximum thickness of subarachnoid blood immediately after nonsurgical resuscitation predicted 1-month mortality with 83.9% sensitivity and 67.1% specificity and that its predictive value was similar to that of the GCS score.^26^ This is important as it may imply that providers should consider the presence of traumatic SAH and explore the predictive value of the maximum thickness of subarachnoid blood as a new independent prognostic marker of mortality. The maximum thickness of subarachnoid blood might be useful as an additional tool for risk stratification and decision-making in the acute phase of traumatic SAH. More research into mortality, clinical and CT progression, neurosurgical intervention, and ICU course is needed.

### Imaging

We hypothesized that there would be more studies on diagnostic and imaging; however, only three studies met the inclusion criteria. We found that repeat outpatient CT of asymptomatic patients after nonoperative cerebral contusion and tSAH is unlikely to demonstrate significant new pathology. Given the cost and radiation exposure associated with CT, it may be that imaging should be reserved for patients with significant symptoms or focal findings on neurological examination. We also found that SWI may be better than CT in detecting intraventricular hemorrhage. SWI is very sensitive to small amounts of SAH. This could be used to help differentiate SAH from veins. However, SWI performs poorly in detecting basilar cistern SAH. Overall, SWI may have the potential to provide complementary information to CT in imaging traumatic SAH.

### Clinical Management and Prognosis

We identified nine studies that discussed the clinical management and prognosis of tSAH. We found that iTSAH, regardless of admission GCS score, is a less severe intracranial injury that is highly unlikely to require aggressive operative, medical, or endovascular intervention. It is also unlikely to be associated with significant neurologic morbidity or mortality, except perhaps in elderly patients.^35^ Age, initial coma scale, extensive tSAH, and IVH are independent predictors of poor outcome in the cohort of tSAH patients. Statistically, patients with extensive tSAH are significantly more likely to have vasospasm.^34^ We found that iTSAH patients with impaired coagulation, especially those with elevated INR, are at risk of clinical and radiologic deterioration. Despite coagulation status, routine repetition of cranial CT scan is recommended in patients with iTSAH to detect potential radiologic progression, which, if detected, should result in close observation.^37^ Patients with traumatic SAH and coexisting types of intracranial hemorrhage have a higher adjusted odds ratio of mortality when compared with patients with an isolated traumatic SAH.^36^ On imaging, the presence of midline tSAH on initial CT was associated with poor early and long-term outcomes, probably due to severe DAI. Conversely, the absence of midline tSAH on initial CT was a reliable marker to exclude severe DAI.^32^ Furthermore, the maximum thickness of traumatic SAH was independently associated with neurological outcome and death but not with the extent or location of hemorrhage.^31^ The amount of subarachnoid blood and associated parenchymal damage are independent factors associated with CT progression, thus linking poor outcomes and CT changes.^33^ Results do not support the finding of a beneficial effect of nimodipine on outcome in patients with traumatic subarachnoid hemorrhage.^30^ One study established an algorithm with three components (head AIS score ≤4, Cr <1.4, and age <76 years) to predict mortality in patients with isolated tSAH. The algorithm may aid in identifying patients with a high risk of poor outcomes.^36^

### Traumatic Aneurysm

Only one study met the inclusion criteria for post aneurysmal tSAH. We found that the mechanism of injury does not correlate with the presence of intracranial aneurysmal disease in patients with tSAH. The presence of central tSAH and intraventricular bleeding may indicate the need for selective performance of follow-up CTA, especially in the simultaneous presence of hypertension. Patients with isolated peripheral tSAH on initial trauma work-up do not seem to warrant subsequent CTA performance.

## Conclusions

TSAH is a public health problem of significant proportions due to the global burden of disease and its disproportionate effect on LMICs. We found that patients with tSAH associated with mTBI have a lower risk of clinical deterioration and surgical intervention. The routine implementation of CT scans, mandatory neurosurgery consultations, and high-intensity observations are not necessary in most cases. Imaging is especially critical for tSAH associated with sTBI. The Stockholm and Helsinki scores may offer improvements in outcome predictions to target intensive care and surgical treatment early on in the course of treatment. The decision tree that evaluates head AIS, Cr, and age may help identify patients with a higher risk of mortality in conjunction with imaging. The presence of central tSAH and intraventricular bleeding may warrant CTA to rule out post-traumatic aneurysmal SAH.

The principal limitation of this and any scoping review is the quality of the included studies. We did not use a tool to assess the quality of the included literature as this is not characteristically a component to scoping review methodology. There were no randomized-controlled trials, leaving the included studies to a higher risk of bias. There are invariably other ways to group the included studies. However, we arranged the studies into five groups based on our best understanding of the evidence.

### Recommendations for research

The evidence on tSAH and mild TBI greatly outweighs that which is available for tSAH and severe TBI. More research is needed for mortality, CT progression, neurosurgical intervention, and ICU course. Furthermore, more research is needed for diagnostic imaging. With advances in imaging occurring at a high rate, patients need to receive optimal care, for which diagnostic capability is a priority. More research is needed to uncover the role of vasospasm in tSAH, as it is a common sequela that is improperly understood.

## Data Availability

All data is available as a supplementary file.

## Author contribution

This manuscript is original work. No part of the manuscript has been published or submitted elsewhere for publication. All authors have read and approved the full version of this manuscript and its submission to this journal. The development of the submitted manuscript has adhered to ethical standards. All authors contributed to the development of the ideas, writing and final review of the submitted manuscript.

## Appendices

### Appendix I: Search strategy

| MEDLINE | Query | Records Retrieved |
| --- | --- | --- |
| #1 | ((("injuries"[MeSH Subheading] OR "injuries"[All Fields] OR "trauma"[All Fields] OR "wounds and injuries"[MeSH Terms] OR ("wounds"[All Fields] AND "injuries"[All Fields]) OR "wounds and injuries"[All Fields] OR "trauma s"[All Fields] OR "traumas"[All Fields]) AND ("subarachnoid haemorrhage"[All Fields] OR "subarachnoid hemorrhage"[MeSH Terms] OR ("subarachnoid"[All Fields] AND "hemorrhage"[All Fields]) OR "subarachnoid hemorrhage"[All Fields]) AND ("diagnosable"[All Fields] OR "diagnosi"[All Fields] OR "diagnosis"[MeSH Terms] OR "diagnosis"[All Fields] OR "diagnose"[All Fields] OR "diagnosed"[All Fields] OR "diagnoses"[All Fields] OR "diagnosing"[All Fields] OR "diagnosis"[MeSH Subheading])) OR ((("injuries"[MeSH Subheading] OR "injuries"[All Fields] OR "trauma"[All Fields] OR "wounds and injuries"[MeSH Terms] OR ("wounds"[All Fields] AND "injuries"[All Fields]) OR "wounds and injuries"[All Fields] OR "trauma s"[All Fields] OR "traumas"[All Fields]) AND ("subarachnoid haemorrhage"[All Fields] OR "subarachnoid hemorrhage"[MeSH Terms] OR ("subarachnoid"[All Fields] AND "hemorrhage"[All Fields]) OR "subarachnoid hemorrhage"[All Fields]) AND ("diagnosable"[All Fields] OR "diagnosi"[All Fields] OR "diagnosis"[MeSH Terms] OR "diagnosis"[All Fields] OR "diagnose"[All Fields] OR "diagnosed"[All Fields] OR "diagnoses"[All Fields] OR "diagnosing"[All Fields] OR "diagnosis"[MeSH Subheading])) | 3,430 |
| Filters | **Humans, English, French, Spanish from 2005 - 2020** | 1,678 |

